# Estimating the number of symptomatic SARS-CoV-2 infections among vaccinated individuals in the United States—January–April, 2021

**DOI:** 10.1101/2021.08.03.21261442

**Authors:** Kiersten J. Kugeler, John Williamson, Aaron T. Curns, Jessica M. Healy, Leisha D. Nolen, Thomas A. Clark, Stacey W. Martin, Marc Fischer

## Abstract

As of March 2021, three COVID-19 vaccines have been authorized by the U.S. Food and Drug Administration (FDA) for use in the United States. Each has substantial efficacy in preventing COVID-19. However, as efficacy from trials was <100% for all three vaccines, disease in vaccinated people is expected to occur. We created a spreadsheet-based tool to estimate the number of symptomatic vaccine breakthrough infections based on published vaccine efficacy (VE) data, percent of the population that has been fully vaccinated, and average number of COVID-19 cases reported per day. We estimate that approximately 51,000 symptomatic vaccine breakthrough infections (95% CI: ∼48,000–55,000 cases) occurred in the United States during January–April 2021 among >77 million fully vaccinated people, reflecting <0.5% of COVID-19 cases that occurred during that time. With ongoing SARS-CoV-2 transmission and increasing numbers of people vaccinated in the United States, vaccine breakthrough infections will continue to accumulate before population immunity is sufficient to interrupt transmission. Understanding expectations regarding number of vaccine breakthrough infections enables accurate public health messaging to help ensure that the occurrence of such cases does not negatively affect vaccine perceptions, confidence, and uptake.

## Introduction

Widespread uptake of safe and effective vaccines is critical to controlling the COVID-19 pandemic. Three COVID-19 vaccines have been authorized by the U.S. Food and Drug Administration (FDA) for use in the United States, including the 2-dose Pfizer-BioNTech BNT162b2 mRNA and Moderna mRNA-1273 vaccines and the single-dose adenovirus-based Johnson & Johnson/Janssen Ad.26.COV2.S vaccine (1-3). In large, randomized controlled trials, the Pfizer-BioNTech and Moderna mRNA vaccines each had an efficacy of ≥94% in preventing symptomatic, laboratory-confirmed COVID-19 following the 2-dose series (4, 5). Among the over 32,000 people who received either the Pfizer-BioNTech or Moderna vaccine during those clinical trials, 20 developed COVID-19 after vaccination. Among the 19,514 people randomized to receive Janssen vaccine during those trials, 116 developed COVID-19 following vaccination, resulting in an efficacy of 67% against moderate-to-severe COVID-19 (6). The Pfizer-BioNTech and Moderna vaccines were authorized by FDA and recommended by the Advisory Committee of Immunization Practices for use in the United States in December 2020 (2, 3, 7, 8), while the Janssen vaccine was authorized and recommended for use at the end of February 2021 (1, 9). Vaccine administration in the United States began within a few days of authorization for each vaccine.

As no vaccine is 100% effective at preventing illness, COVID-19 occurring among vaccinated people, or symptomatic vaccine breakthrough infections, are expected. Amid increases in vaccination coverage in the setting of widespread SARS-CoV-2 transmission, the numbers of vaccine breakthrough infections could be substantial. We estimated the number of symptomatic vaccine breakthrough infections expected in the United States based on published vaccine efficacy (VE) data, percent of the population that had been fully vaccinated, and reported COVID-19 case counts.

## Methods

We developed a tool in Microsoft Excel^©^ to estimate the expected number of symptomatic vaccine breakthrough infections per day among U.S. adults (aged ≥18 years). Inputs are the 7- day moving average for daily numbers of COVID-19 cases in the United States as reported to the Centers for Disease Control and Prevention (CDC), the cumulative number of persons fully vaccinated as reported to CDC as of 14 days prior to each 7-day average case count, and VE data from phase 3 trials of the three vaccines authorized in the United States (1-3, 10, 11). The number of symptomatic vaccine breakthrough infections, rather than all symptomatic and asymptomatic vaccine breakthrough infections, were calculated because prevention of symptomatic disease was the primary phase 3 clinical trial endpoint reported for the COVID-19 vaccines authorized in the United States.

Breakthrough cases were defined as those occurring in persons ≥14 days after completion of vaccination with an authorized COVID-19 vaccine, a delay to reflect when maximum immunity conveyed by vaccination is reached. Given the similar reported VE from clinical trials for the Pfizer-BioNTech and Moderna vaccines, the average VE of 94.6% was used for both mRNA vaccines in the calculator (4, 5), while 66.9% VE was used for the Janssen vaccine (1). Calculations were restricted to persons aged ≥18 years, the primary population that received vaccines during the study period. Because available data suggest that 88.3% of reported U.S. cases occur among persons ≥18 years of age, we similarly restricted the denominator data used for this analysis to 88.3% of the total population per 2019 U.S. census estimates (12).

The number of symptomatic vaccine breakthrough infections expected per day is a function of VE and vaccination coverage in the population. For these calculations, *C* denotes the 7-day moving average for daily number of reported COVID-19 cases and *V* represents different vaccination “groups” according to numeric subscripts: 0 for unvaccinated or not fully vaccinated, 1 for Janssen vaccine, and 2 for Moderna and Pfizer-BioNTech vaccines; *%V* is the percent of the population fully vaccinated in each vaccine group. VE is calculated as (1 – the risk ratio [RR]), where RR is the ratio of confirmed symptomatic SARS-CoV-2 infections per 1000 person-years among those receiving vaccine in phase 3 trials divided by those receiving placebo. The Janssen RR_1_ was 0.331 (VE_1_ = 0.669), the Pfizer-BioNTech and Moderna RR_2_ was 0.054 (VE_2_ = 0.946), while RR_0_ was defined as 1 (VE_0_ = 0) for people who are unvaccinated or not fully vaccinated. The expected number of symptomatic vaccine breakthrough infections per day is calculated as:

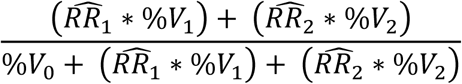

Variance was calculated based on available phase 3 clinical trial data for the Pfizer-BioNTech and Moderna vaccine trials using Poisson regression models. The pooled variance of the expected symptomatic vaccine breakthrough infections was estimated to be 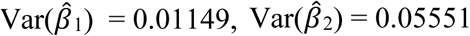, with 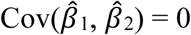 and calculated as

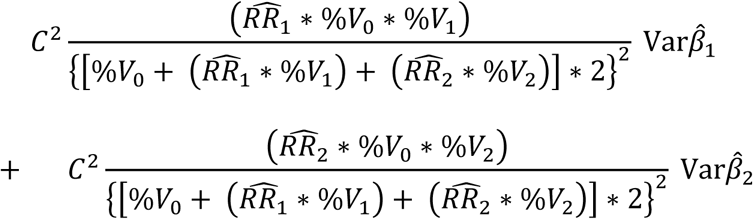

The first persons in the United States to be vaccinated against SARS-CoV-2 completed their 2-dose series during the week of January 4, 2021. Therefore, we began calculating the expected number of symptomatic vaccine breakthrough infections 14 days later, the week beginning January 17. We calculated weekly estimates using average case counts and vaccination coverage as of the Sunday beginning each week and then multiplied the daily estimate by seven. We estimated per-week symptomatic breakthrough infections through the week beginning April 25. Incorporation of Janssen vaccine into the estimates began in mid-March. We calculated cumulative expected counts to date by summing weekly expected vaccine breakthrough case counts. We derived 95% confidence intervals (CI) around cumulative counts by summing weekly variances as described above into standard CI calculations.

To understand the relative influence of community transmission, vaccination coverage, and VE in determining the number of expected vaccine breakthrough infections, we calculated expected cumulative counts during the same time period under three hypothetical scenarios: 1) doubling the average daily COVID-19 case counts each week; 2) doubling cumulative vaccination coverage each week; and 3) modifying population vaccination coverage during January–April such that it entirely reflected VE of 67%, VE associated with the Janssen vaccine.

## Results

Over 11 million COVID-19 cases were reported in the United States during January–April 2021 (10). The number of COVID-19 cases reported per day during this period ranged from a high of approximately 217,000 cases in January to a low of approximately 53,000 cases in March (10). The estimated number of symptomatic vaccine breakthrough infections in the United States ranged from a low of approximately nine per week in January to 11,300 during the last week of April (Table 1 and Figure).

**Table 1.**
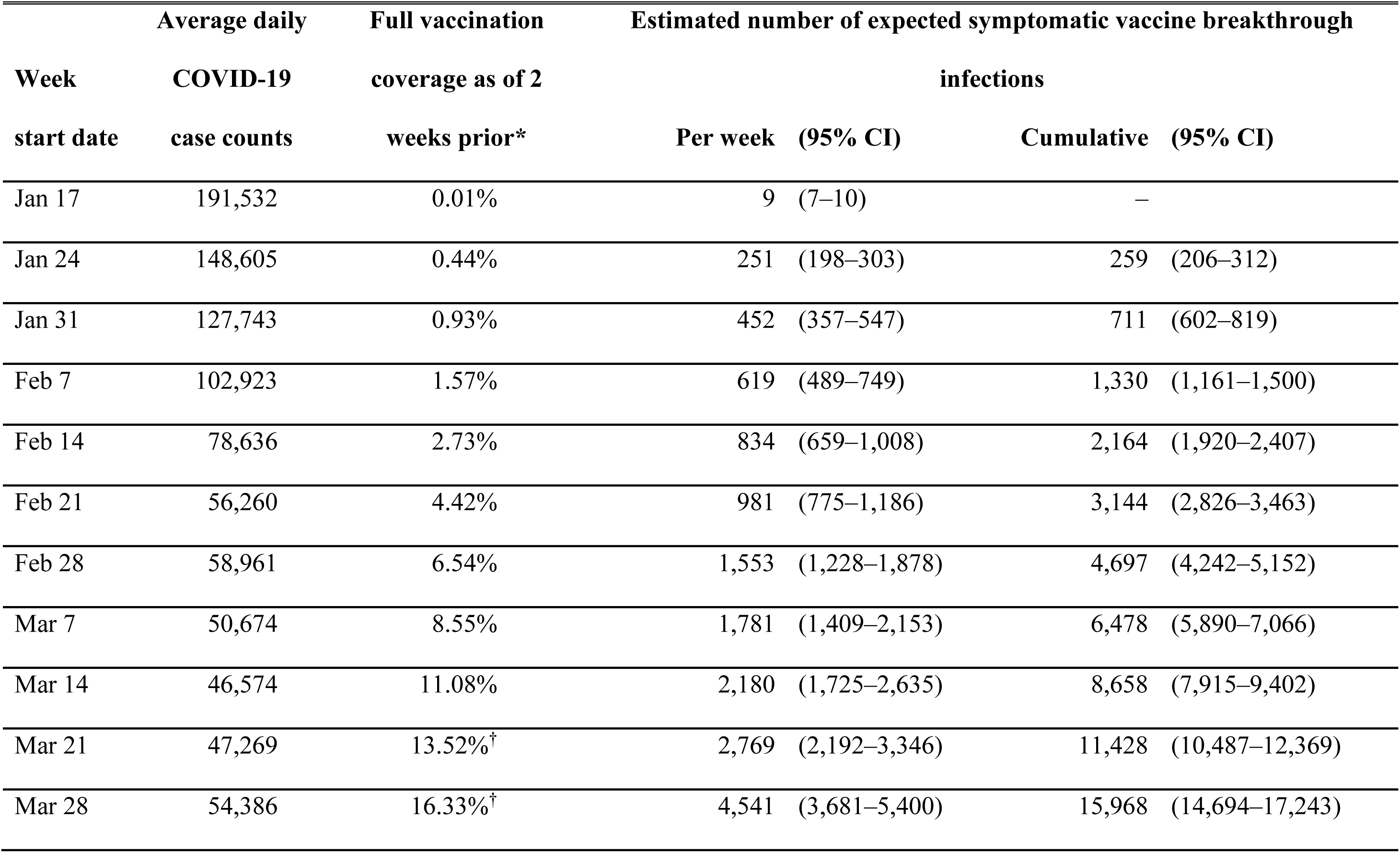
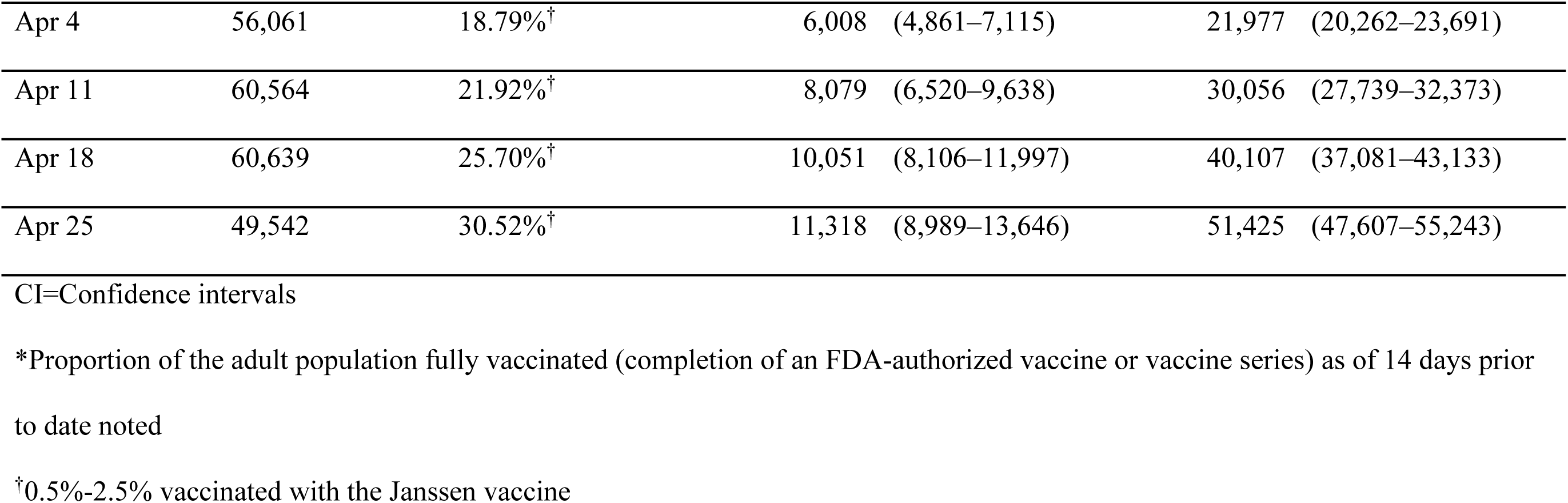
COVID-19 case counts, vaccine coverage, and expected number of symptomatic vaccine breakthrough infections among adults (aged ≥18 years), by week — United States, January–April 2021.

**Figure.**
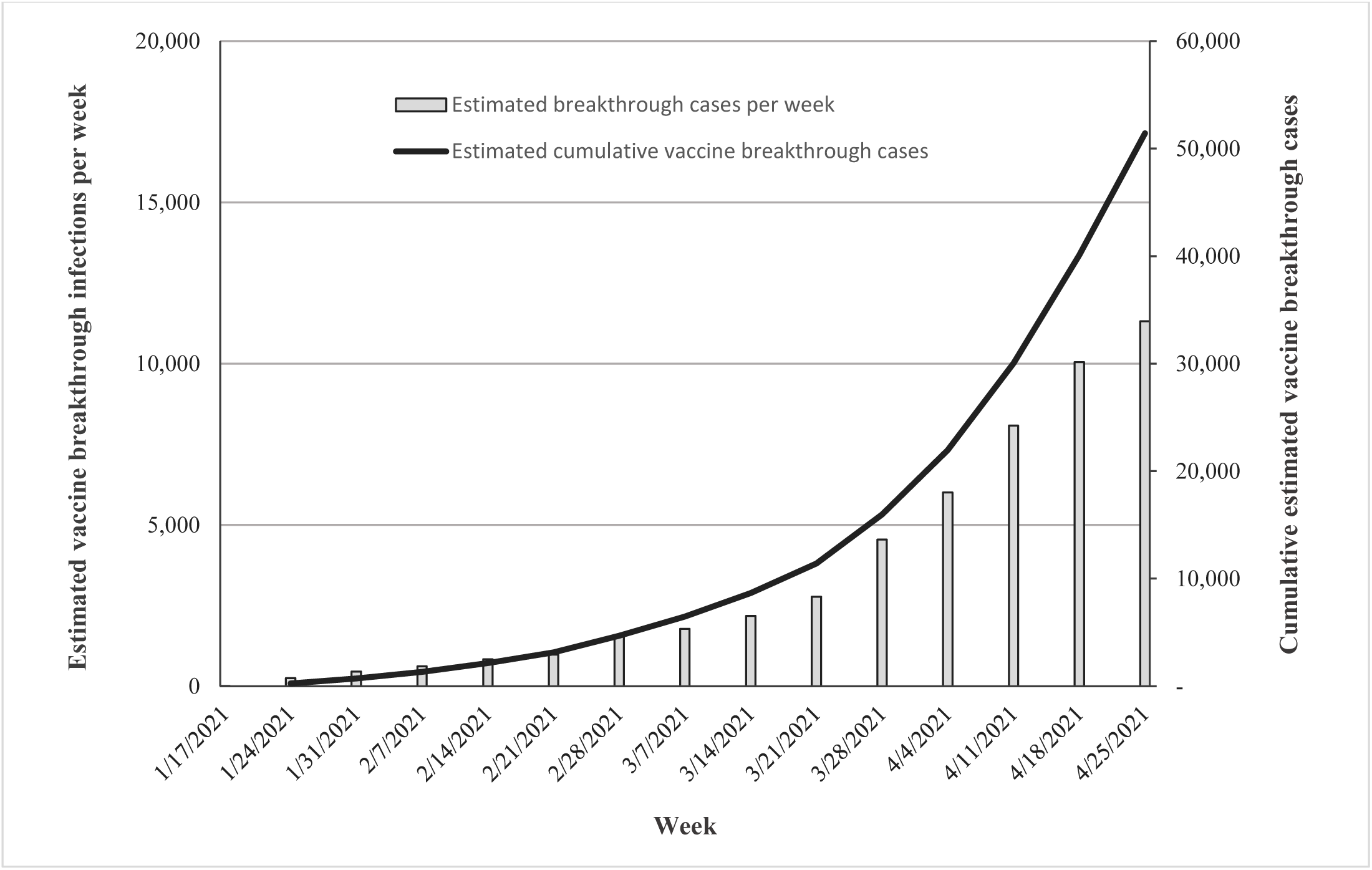
Estimated symptomatic COVID-19 vaccine breakthrough infections per week and cumulatively in the United States from January–April 2021.

As of the end of April, we estimate that a total of 51,425 symptomatic vaccine breakthrough infections (95% CI: 47,607–55,243 cases) occurred in the United States among >77 million fully vaccinated people (Table 1 and Figure). On average, starting in February, the number of expected vaccine breakthrough infections increased by 37% each week, becoming exponential; this trajectory slowed during the last week of April amid falling numbers of COVID-19 cases in the United States (Figure) (10). The number of expected vaccine breakthrough infections during that time translates to a cumulative incidence of approximately 66 vaccine breakthrough infections per 100,000 fully vaccinated people.

The expected number of vaccine breakthrough infections varied substantially under different hypothetical scenarios reflective of 1) doubling daily average case counts, 2) doubling the vaccination coverage, and 3) all vaccination occurring at VE of 67% (Table 2). The relationship between COVID-19 cases and the expected number of vaccine breakthrough infections was proportional (i.e., when case counts doubled, so did symptomatic vaccine breakthrough infections). The doubling of vaccination coverage more than doubled the number of expected breakthrough cases. VE had the most influence on the expected number of symptomatic vaccine breakthrough infections. Compared to vaccination with an average VE of nearly 95%, as occurred during January through April in the United States, a hypothetical scenario in which all vaccination occurred at VE of about 67% nearly quadrupled the number of expected symptomatic vaccine breakthrough infections without modifying other parameters.

**Table 2.**
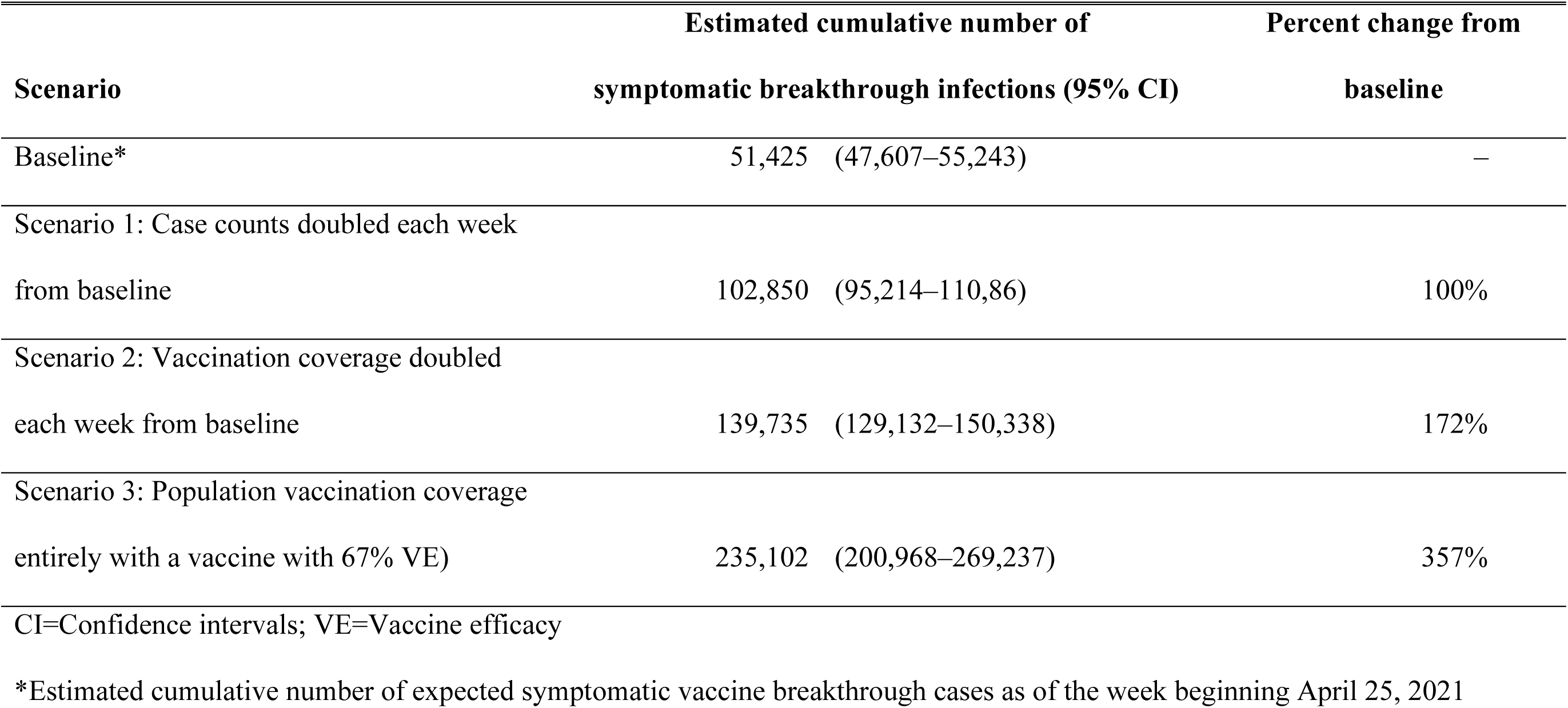
Expected cumulative number of symptomatic COVID-19 vaccine breakthrough infections under differing hypothetical scenarios of disease incidence, vaccination coverage, and vaccine efficacy—United States, January–April 2021.

| Scenario | Estimated cumulative number of symptomatic breakthrough infections (95% CI) | Percent change from baseline |
| --- | --- | --- |
| Baseline* | 51,425 (47,607–55,243) | – |
| Scenario 1: Case counts doubled each week from baseline | 102,850 (95,214–110,86) | 100% |
| Scenario 2: Vaccination coverage doubled each week from baseline | 139,735 (129,132–150,338) | 172% |
| Scenario 3: Population vaccination coverage entirely with a vaccine with 67% VE) | 235,102 (200,968–269,237) | 357% |
| CI=Confidence intervals; VE=Vaccine efficacy |  |  |

\*Estimated cumulative number of expected symptomatic vaccine breakthrough cases as of the week beginning April 25, 2021

## Discussion

We created a spreadsheet-based calculator to estimate the number of symptomatic vaccine breakthrough infections in the United States based on the average number of COVID-19 cases, percent of the population fully vaccinated, and published efficacies of the three FDA-authorized vaccines. Using this tool, we estimate that approximately 51,000 symptomatic SARS-CoV-2 vaccine breakthrough infections occurred among the over 77 million people fully vaccinated in the United States by the middle of April. These symptomatic breakthrough infections represent <0.5% of all COVID-19 cases that occurred during January–April 2021. Vaccine breakthrough infections occurred in only a small fraction of all vaccinated persons and comprised only a small fraction of all COVID-19 cases. Understanding the number of expected vaccine breakthrough infections is important for accurate public health messaging to help ensure that the occurrence of such cases does not negatively affect vaccine perceptions, confidence, and uptake.

We developed this tool incorporating ideal VE scenarios. Real-world vaccine effectiveness may be lower, particularly for people who are older or have underlying health conditions (13, 14). Lower average effectiveness also could result from decreased protection against certain SARS-CoV-2 variants. Nevertheless, early vaccine effectiveness studies in the United States and elsewhere demonstrate high effectiveness of mRNA vaccines against symptomatic infection and severe disease in various real-world situations, including amid increasing circulation of SARS-CoV-2 variants of concern (15, 16), including approximately 95% effectiveness among large cohorts of healthcare workers and >85% among residents of skilled nursing facilities (17-22). While we used efficacy data from clinical trials as inputs to estimate expected vaccine breakthrough infections, these inputs could be updated using real-world vaccine effectiveness data as those become increasingly available.

COVID-19 vaccines have demonstrated the ability to mitigate risk of severe disease, hospitalization, and death among persons infected following vaccination (4, 5, 13, 17). Clinical trial endpoints utilized here were for prevention of symptomatic infection; the effectiveness of authorized vaccines at preventing asymptomatic SARS-CoV-2 infection is still unclear but preliminary reports suggest the mRNA vaccines may be >90% effective at preventing infection (13, 14, 17, 21, 23). We did not incorporate asymptomatic infections into these calculations.

The analytic approach described here is based on several assumptions and limitations that affect how our results should be interpreted. First, this approach is based on reported case counts and does not account for the population at-risk, susceptibility of persons previously infected, or duration of immunity following vaccination. Second, these calculations assume that vaccinated and unvaccinated people have the same risk of exposure to SARS-CoV-2, which may not be true at the population level. Third, we define vaccine breakthrough infections as those occurring more than two weeks after completion of vaccination; these figures do not include people who may become infected following partial vaccination or prior to 14 days following completion of vaccination. Fourth, reported COVID-19 case counts stratified by patient age are not available from all states (12). Our assumption that adults comprise 88.3% of reported cases reflects cumulative trends since the beginning of the pandemic; these data may not reflect the age distribution during the weeks included here and only approximate the number of COVID-19 cases occurring among adults. If the proportion of total cases occurring among adults decreased over time, our assumption would yield an overestimate of vaccine breakthrough cases. As younger age groups commence vaccination, calculations should be updated to reflect the entire population. Lastly, reporting delays among both COVID-19 case and vaccine administration data vary, and data are often updated retrospectively. Therefore, the figures used for these calculations should be viewed as approximations. Collectively, because of these limitations, the specific estimated counts should be interpreted with caution. Nevertheless, they provide useful context for guiding expectations.

Risk reduction provided by any vaccination is inherently relative, and the number of cases among vaccinated persons assuming equal exposure as unvaccinated persons is directly linked to vaccination coverage and disease incidence. Even with highly effective vaccines, given the large number of people being vaccinated in the United States and high levels of SARS-CoV- 2 transmission in some parts of the country, tens of thousands of symptomatic vaccine breakthrough infections are expected. The methods described here can be used by public health officials to determine if the frequency of vaccine breakthrough infections reported in their jurisdictions are consistent with expectations based on vaccine efficacy from clinical trials. Furthermore, public health messaging regarding expected vaccine breakthrough infections is important to assure the public that this is expected, is not cause for alarm, and does not indicate that vaccines are not preventing COVID-19.

Vaccine breakthrough infections are expected to continue to accumulate amid ongoing widespread community transmission of SARS-CoV-2 and increasing vaccination coverage. However, the number of COVID-19 cases, hospitalizations, and deaths prevented among vaccinated persons will far exceed the numbers of vaccine breakthrough infections. CDC is working with public health officials nationwide to monitor vaccine breakthrough infections, including those that result in severe disease, and to identify unexpected trends in characteristics of people with vaccine breakthrough infections or patterns associated with infecting strains.

## Data Availability

All input data are publicly available

## Acknowledgments

CDC’s COVID-19 Vaccine Breakthrough Case Investigation Team.

## Funding

This work was supported by the U.S. Centers for Disease Control and Prevention.

## Disclaimers

The findings and conclusions in this report are those of the author(s) and do not necessarily represent the official position of the Centers for Disease Control and Prevention. Names of specific vendors, manufacturers, or products are included for informational purposes and does not imply endorsement of the vendors, manufacturers, or products by the Centers for Disease Control and Prevention or the U.S Department of Health and Human Services.

## Notes

### Competing Interest Statement

The authors have declared no competing interest.

### Funding Statement

No external funding was received.

### Author Declarations

This activity was conducted consistent with applicable federal policy.

